# Resilience and green spaces: association with stress among contact centre agents in the Philippines

**DOI:** 10.1101/2020.06.14.20131276

**Authors:** Celine P. Villanueva, Richard Braulio J. Labao, Katherine Regine Anne G. Tran, Nathaniel Reihann B. Gonzalez, Joseph M. Luna, Kristine Mallory R. Ochava, Catherine M. Capio

**Author notes:** **Correspondence** Dr. Catherine M. Capio, Centre for Educational and Developmental Sciences, The Education University of Hong Kong, Hong Kong SAR,.

## Abstract

The work environment in Philippine contact centres had been shown to expose employees to factors that contribute to work-related stress; health promotion strategies that could mitigate the impacts are needed. With the framework that stress is experienced as a result of the interactions of an individual with the environment, this study examined the relationship of stress with individual resilience and the presence of urban green spaces (UGS) in the environment. The study involved employees (N = 186) from six contact centres in the capital region of the Philippines, where a large number of contact centre service providers are located. A two-stage survey was administered online using standardised instruments to measure stress (10-item Perceived Stress Scale) and resilience (Connor-Davidson Resilience Scale), customised questions to gather demographic information, and open-ended questions to probe on participants’ insights. Google Earth Pro was used to conduct satellite mapping of UGS, followed by on-site ocular inspection. This study revealed that participants’ average stress level was categorised as high. The percentages of UGS in the vicinity of the study sites were categorised as low. Linear regression revealed that amongst individual and environment factors, resilience, household income, and awareness of UGS in the vicinity were significant predictors of the participants’ stress levels. Health promotion in Philippine contact centres could consider strategies that include resilience building, enhancing income security, and promoting the awareness of UGS within the workplace vicinity.

## INTRODUCTION

Advances in information and communication technologies have accelerated globalisation as workplaces have been enabled to overcome time differences and space constraints (Min et al., 2019). This is most evident in business process outsourcing (BPO) in which segments of operations are subcontracted to external service providers that are not necessarily in the same country or region (Duening and Click, 2005). As such, most cases entail shift work that follows a time zone that is different from that of the workers (i.e., increase in night shifts). The Philippines had emerged as one of the leading international BPO providers, particularly for administrative services (Duening and Click, 2005). A large portion of the Philippine BPO industry is made up of the sub-sector of contact centres, which “provide both inbound and outbound voice services for sales, customer service, technical support and other business processes” (Errighi et al., 2016, p.2). Despite the low skill level needed for contact centres, the sub-sector attracts a large number of workers, including high-skilled professionals, because of wages that are above the national average (Amante, 2010). However, the International Labour Organization (ILO) identified BPO employees as a population that is exposed to stressful environments and heightened discontent, which had been evident in high employee turnover rates (Errighi et al., 2016).

Stress had been defined as “a particular relationship between the person and the environment that is appraised by the person as taxing or exceeding his or her resources and endangering his or her wellbeing” (Lazarus and Folkman, 1984, p.19). The environment of Philippine contact centres had been described as stress-inducing due to task-related factors. For instance, high volumes of work need to be completed within limited amounts of time, while voice calls were closely monitored to ensure that workers meet pre-determined daily quotas (Amante, 2010, Hechanova, 2013). These situations appear to compromise contact centre agents’ wellbeing. It had been reported that conflicts arising from irate customers, along with unpredictable work schedules contribute to workers’ disturbed sleeping patterns. Both physical and psychosocial symptoms including, but not limited to, muscular tension, insomnia, and headaches had also been attributed to stress experienced in the contact centre environment (Amante, 2010, Lozano-Kühne et al., 2012). It is not surprising that stress had been considered the leading cause for employee turnover (Hechanova, 2013).

Faced with the challenges of a stressful working environment, employees might still maintain an ability to adapt and maintain psychological stability and focus, which has been referred to as resilience (Shatte, 2012). It has also been described as an individual’s ability to respond well to stressors with reduced harmful consequences, and had been shown to have a protective effect for workers in highly stressful environments (Shatte et al., 2017). Resilience and its potential to influence workers’ stress levels has yet to be explored in contact centre settings. In the Philippines, resilience had been examined in the context of disasters (e.g., Tanyag, 2018, Usamah et al., 2014) but not in the perspective of promoting health in the workplace. While the value of resilience training in the workplace had become increasingly evident (Robertson et al., 2015), there is a gap in our understanding of how individual resilience interacts with work-related stress in Philippine contact centres.

Looking at the environment, there has been growing interest in the health benefits associated with urban green spaces (UGS) which include stress relief (WHO, 2016). UGS has been described broadly as any green space, public open space, or park in an urban setting (Lee et al., 2015), which might include sports fields, natural meadows, wetlands, and other ecosystems that facilitate physical activity and relaxation (WHO, 2016). In this study, UGS refers to accessible and open areas of greenery excluding those designated as pathways amongst urban buildings. Studies across the globe have suggested that exposure to UGS is positively correlated to multi-dimensional aspects of wellbeing (WHO, 2016). For instance, UGS had been linked to longevity among senior citizens who either used green spaces or had green spaces near their residences (Takano et al., 2002). It had been argued that UGS provide a venue for activities that benefit the wellbeing of people, as it became evident that the presence of green spaces contributed to increasing the frequency of physical activity among adults (Astell-Burt et al., 2014). Individuals with a higher percentage of greenery around them also felt less lonely and perceived greater social support (Maas et al., 2009), while high quality green spaces had been linked to an improved sense of social connectedness (de Vries et al., 2013).

Contact centre agents in the Philippines make up a large workforce who are exposed to factors that contribute to work-related stress, and health promotion strategies that could mitigate the impact are needed. Based on the framework that stress depends on the interactions of individual and environment characteristics (Lazarus and Folkman, 1984), evidence suggests that resilience and UGS present potential factors to consider. Given limited context-specific research, this study examined the relationship of stress with environment (i.e., UGS) and individual (i.e., resilience) factors amongst a sample of contact centre employees in an urban region of the Philippines. Based on data gathered through standardised measurement instruments, it was hypothesised that stress will have a negative association with resilience and presence of UGS.

## MATERIALS AND METHODS

This is a cross-sectional study using a concurrent mixed-methods research design with the quantitative component being the main focus (QUAN + qual) (Schoonenboom and Johnson, 2017). All procedures were approved by the institutional ethics review board, and participants provided informed consent.

### Participants

Employees from six contact centres within the national capital region (NCR) of the Philippines participated in the study. Eligible participants were those who were (a) working full-time for at least 6 months at the time of recruitment, and (b) and at least 18 years old. Those who were currently prescribed with medications for psychiatric symptoms were excluded. Out of 202 potential participants, 94% (N = 190) completed the first questionnaire that measured their perceived levels of stress. Four individuals were excluded after the first survey indicated extreme levels of perceived stress (see Methods section for the cut-off score); they were subsequently advised to seek professional health consultation. Out of the 186 participants who were invited to respond to the second questionnaire that measured resilience and obtained UGS-related information, 48% (n = 89) submitted complete responses; the others submitted partial responses which were not included in the subsequent analysis. The final sample consisted of 30.3% males and 69.7% females. The participants’ age ranged from 19 to 53 years (*M* = 30, *SD* = 7.47).

### Procedures

#### Questionnaires

Two sets of questionnaires were administered in sequence using an online platform. Survey links were shared to contact centre representatives, who then distributed them to the employees who met the eligibility criteria.

In the first questionnaire, perceived stress was measured using the 10-item Perceived Stress Scale (PSS-10; Cohen et al., 1983) which is one of the most widely-used measures of self-reported stress. It had been shown to be a valid instrument and is not affected by gender bias (Taylor, 2015). It had been translated and validated in multiple sub-groups across the globe (e.g., Al-Dubai et al., 2014, Andreou et al., 2011, Huang et al., 2020). In this study, the original version was deemed appropriate as English is an official language used in Philippine corporate environments.

Using a 5-point Likert scale (0 to 4), PSS-10 measures the degree to which current life situations are deemed stressful. Higher scores indicate higher levels of perceived stress and the highest possible score is 40. The scores correspond to stress levels as follows: 0-7 as very low, 8-11 as low, 12-15 as average, 16-20 as high, and ≥21 as very high; those who score >31 are deemed to be in extreme stress (Roe et al., 2017). In this study, participants who scored greater than 31 were advised to seek professional health consultation and were not invited to proceed to the second questionnaire.

The second questionnaire included the 10-item Connor-Davidson Resilience Scale (CD-RISC; Connor and Davidson, 2003) which had been shown to be a valid measure of resilience. Higher scores in CD-RISC indicate greater resilience capability. The questionnaire also sought to obtain other individual information that included age, household income category, and frequency of engagement in exercise. It included adapted items on UGS interactions (Lottrup et al., 2013) which used a 5-point Likert scale to assess visibility (1 = very noticeable, 5 = no green environment) and physical accessibility (1 = accessible and used often, 5 = none that is accessible) of UGS in relation to the participants’ workplace. A dichotomous item (Yes/No) assessed the participants’ awareness of UGS nearby. Finally, the questionnaire included open-ended questions to probe on their insights in relation to work-related stress and UGS.

### UGS Measurement

Google Earth Pro had been recommended and used for satellite mapping of urban spaces (Taylor and Lovell, 2012), and was used in this study to identify the UGS area proximal to the sites of the six contact centres. The study areas were defined at a 300-meter radius, generating a 282,600 square-meter area around the workplace. Ocular inspections were subsequently conducted by researchers who physically visited the sites to verify and confirm the UGS areas that were identified in the satellite mapping process. The area of identified green spaces was used to calculate the percentage of UGS (%UGS) within the area using the following formula:

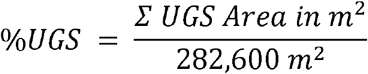

Area-based measurement is the most frequently used approach to quantify exposure to UGS (Kondo et al., 2018). The WHO (2016) also recommends the use of proximity-based indicators, one of which is UGS percentage (%UGS) of the area. The proportion of urban green space within a person’s environment combines both proximity and quantity. Thompson et al. (2012) classified %UGS into four categories: low (<23%), medium-low (24%-34%), medium-high (35-46%), and high (>46%).

### Data Analyses

A dependent mixed-methods analysis approach was used (Schoonenboom and Johnson, 2017), such that quantitative data analyses were first performed followed by qualitative analysis to follow-up on the findings of the quantitative component. Descriptive and inferential statistics were performed to identify the relationship of perceived stress levels with resilience, UGS, and other individual characteristics. All statistical tests were performed using SPSS 25, and statistical significance was set at *p* < 0.05. Thematic content analysis was performed to examine the qualitative data.

## RESULTS

### Participants’ characteristics

Out of 190 respondents who completed the first questionnaire, 104 (55%) were categorized to have high or very high stress levels. The mean PSS score was 17.45 (SD=6.09). Four (2%) respondents were found to have scores that are greater than 31, and were categorized to have extremely high perceived stress. These respondents were advised to seek professional health consultation and were not invited to the second questionnaire as a precautionary measure.

From 89 participants who completed the second questionnaire, the mean PSS and CD-RISC scores were 17.26 (*SD* = 5.32) and 32.49 (*SD* = 4.86) respectively. Majority (61.8%) of the participants reported household incomes in categories that were above the national poverty line (Philippine Statistics Authority, 2019). A relatively small proportion of the participants exercised every day (12.4%), while a greater number did not exercise at all (31.5%); the median rating for exercise frequency corresponded to “a few times per month”.

### UGS

The mean %UGS measured through satellite mapping for the sites of the six contact centres was 4.96% (*SD* = 5.10). The %UGS for each site is presented in Table 1, and a detailed profile of the surveyed sites is available in the supplementary file. While nearly half of the respondents noted that UGS are noticeable (20.2%) or visible (24.7%), the median rating for UGS visibility was 3 which corresponds to “visible after careful observation”. Majority of the respondents (71.9%) were aware of UGS in the area of their workplaces.

**Table 1.** Total area of UGS and %UGS per study site

| Contact Centre | Total UGS Area (sq.m.) | %UGS |
| --- | --- | --- |
| 1 | 9975.85 | 3.53% |
| 2 | 53,698.33 | 19.00% |
| 3 | 27,255.21 | 9.64% |
| 4 | 0 | 0% |
| 5 | 1999.01 | 0.71% |
| 6 | 10,850.60 | 3.84% |

### Factors associated with stress

Pearson’s product moment correlation test showed a significant negative association between PSS and CD-RISC scores (*r* = −0.455, *p* < 0.001). Respondents with high stress levels reported lower resilience. A significant negative association was also found between PSS scores and age (*r* = −0.251, *p* = 0.018). Older contact agents reported lower perceived stress levels than younger contact agents. The difference in PSS scores between males and females was not significant (*t*(87) = −0.997, *p* = 0.322).

Spearman’s rank correlation coefficient test showed a significant negative association between PSS scores and household income range (*r* = −0.260, *p* = 0.014). Those with higher income tended to have lower stress levels. A significant negative association was also found between PSS scores and frequency of exercise (*r* = −0.246, *<u>p</u>* = 0.020). Respondents who exercised more frequently (*r* = 0.289, *p* = 0.003) reported lower perceived stress levels.

PSS scores were not associated with %UGS, visibility, accessibility, and frequency of use. Independent samples t-test showed that participants who were aware of UGS in the vicinity of their contact centre site reported lower stress levels than those who were not aware (*t*(87) = - 2.737, *p* = 0.008). The mean PSS score for those who were aware of UGS was 16.33 (SD = 4.989), and for those who were not aware of UGS was 19.64 (SD = 5.484).

Stepwise linear regression, considering the significant associated factors (i.e., CD-RISC, age, household income, exercise frequency, UGS awareness), revealed that the best model (*F*(3, 85) = 14.120, *p* < 0.001) included CD-RISC score, household income, and UGS awareness as significant predictors of PSS score (*R*2 = 0.333). Table 2 shows the coefficients of the predictors that were included in the regression.

**Table 2.** Standardised coefficients of the factors tested in the linear regression with participants’ stress levels (PSS scores) as the dependent variable.

| Predictor | Standardised coefficient (Beta) | Sig. |
| --- | --- | --- |
| CD-RISC score | -0.461 | < 0.001 |
| UGS awareness | 0.255 | 0.005 |
| Household income | 0.212 | 0.020 |
| Age | -0.114 | 0.219 |
| Frequency of exercise | 0.158 | 0.087 |

### Qualitative findings

Following a QUAN + qual design, content analysis revealed that the participants identified clients as the primary source of stress at work, particularly those who are incensed and demanding. Stress at work was perceived to produce negative effects on other aspects of participants’ lives, such as their social relationships and personal wellbeing. Emotional instability, in particular, appeared to be the most frequently reported effect of work-related stress. A number of respondents found themselves more irate or depressed when they felt that the stress at work was escalating.

To cope with work-related stress, most respondents reported emotion-focused strategies. Responses under this category include strategies such as positive-thinking and active efforts to keep calm in the presence of stressors. While the employers offer various programs and facilities for wellbeing (including exercise), these did not appear to be perceived as means to cope with stress. In particular, the respondents noted that participation in programs are frequently presented as a requirement, and therefore often perceived as additional work demands rather than opportunities to improve wellbeing. The responses indicated that whilst stress levels are high, there is a greater concern for overall employee conditions and most respondents noted that increased incentives are needed.

Relationships between employees and their supervisors were reportedly positive, and in some cases, even familial. However, none of the respondents associated such interpersonal relationships with mitigating work-related stress. Instead, personal family relationships and friendships were relied upon for social coping (e.g., spending time with family members). Despite stressors being consistently present at work, the respondents reported that they remain in the contact centre industry because of perceived financial benefits. Besides the competitive salary, benefits (e.g. reward packages, health insurance) were noted as motivators to stay in their positions despite work-related stress.

The most frequently reported characterisation of UGS by participants were “dense greenery” and “fresh air”. A few others considered notions of water and beaches. Among those who were aware of UGS around the area of their workplace, no one specifically linked them to coping with stress at work. Nevertheless, the most frequently reported UGS functions are “relaxation spots” and “resting areas”. Majority of the respondents reported that greenery evoked feelings of relaxation. Other activities in UGS that respondents noted include exercising, smoking, and eating.

## DISCUSSION

The work environment of BPOs, especially contact centres in the Philippines, have been shown to be rife with factors that expose employees to stress (Amante, 2010, Errighi et al., 2016, Hechanova, 2013). This current study offers evidence that indeed, contact centre employees in the country continue to experience high levels of stress. A score of 17 and above on the PSS had been categorised as indicative of a high stress level (Roe et al., 2017); the average PSS score of the study sample was higher than this cut-off score, and more than half of the respondents reported high to very high levels of stress. Qualitative findings showed that the participants also noted how their stress levels negatively impact their social relationships and emotional stability. In the context of a relatively low-income and developing country, work-related stress could potentially combine with personal constraints that cascade into health problems. This combination is illustrated in the current findings that those with lower household incomes tended to have higher stress levels; consequently, being at a greater risk of further health issues.

Given that stress is experienced through a process that involves the reciprocal interactions between the stressors in the environment and the responses of the individual (Lazarus and Folkman, 1984), potential health problems might be mitigated through a better understanding of both the environmental and individual factors. Age is one such individual factor, and contact centre practitioners have associated youth with greater resistance to work-related stress (Dharamdass and Fernando, 2017). On the contrary, this current study revealed that higher stress levels were found in younger respondents. Other research had similarly shown younger workers to be more susceptible to stress and less effective with coping (Archer et al., 2015, Siu et al., 2001). This can be understood along the lines of the emotional development theory, which suggests an increase in adaptive coping mechanisms as one ages (Hertel et al., 2013). The ability to adapt could also be apparent through another individual factor – resilience. Indeed, this current study revealed that those who were assessed to have greater resilience reported lower stress levels, independent of age. Resilience is an individual trait but evidence had suggested that attributes of resilience (e.g., emotion regulation, impulse control) could be enhanced and developed (Shatte, 2012). A review had shown that resilience training interventions improve individual ability to adapt and enhance psychosocial wellbeing (Sarkar and Fletcher, 2017). Contact centres could consider trainings to promote resilience among their employees.

In this study, physical activity was assessed in terms of self-reported frequency of exercise. Respondents who exercised more frequently reported lower stress levels. Other studies have shown a similar relationship between physical activity and stress levels (e.g., Branas et al., 2011, Neves et al., 2014). This association could be viewed from a physiologic perspective as exercise is known to stimulate the release of endorphins in the body. Endorphins generate feelings of euphoria and analgesia, and thereby induces a sense of self-efficacy or elevates the mood (Mikkelsen et al., 2017) which counters stressors. However, the qualitative component of this study revealed that the respondents did not seem to consider exercise as a strategy to cope with stress (i.e., only one participant specifically identified exercise as a coping mechanism). This suggests that the contact centre agents in this study might have limited awareness of the mitigating effects of physical activity on stress and its positive relationship with mental wellbeing; deliberate strategies in workplace health promotion could address this limited awareness. It is important, however, that such health promotion strategies not be presented as requirements that workers might perceive as additional burden.

UGS represents an environment factor that could influence work-related stress. While previous studies from developed countries have shown that the presence of UGS contributes to reduction of stress (e.g., Maas et al., 2009, Thompson et al., 2012), this current study showed no association between individual stress levels and objectively measured UGS within a 300-meter radius of the workplace. It is noticeable that UGS was found to be generally sparse around the study sites. UGS that fall below 23% of the measured area are considered low (Thompson et al., 2012), and the measurements reported in this current study ranged from nil to 19% (see Table 1). It is possible that the low UGS measured across the study sites comprised a data variance that was too limited to reveal significant associations with stress levels. The low UGS finding in itself needs to be noted, as it suggests that the urban areas in the vicinity of contact centre offices in the capital of the Philippines tend to be designed with limited allocation for accessible and open green spaces.

Green spaces in urban planning and design are generally within the mandate of public governance and private builders (Bulkeley et al., 2011, Kabisch, 2015). The current study findings, therefore, have implications that need to be addressed through policy and investment. Whilst these aspects are beyond the direct influence of contact centre managements, UGS as an environment factor that could mitigate stress is a health promotion strategy that contact centres could still consider. The findings of this study revealed that participants who were aware, tended to have lower stress levels, compared to those who were not aware of UGS in their vicinity. It is therefore worth considering whether there is value in simply making the employees aware when UGS is within the vicinity of the workplace. The qualitative component of this study showed that participants’ ideas of UGS ranged from dense greenery and fresh air, to beaches and water. One aspect of enhancing workers’ UGS awareness could be promoting a better definition of UGS in the context of workplaces (i.e., open and accessible green spaces). This could work because natural settings tend to draw attention such that by captivating an onlooker and requiring little cognitive effort, executive functions are focused away from stressors and negative emotions (Berto, 2014).

This study offers evidence that could support stress management and health promotion strategies in contact centres in the capital region of the Philippines and comparable territories. To obtain reliable and objective data, standardised approaches were taken to measure stress levels, resilience, and the percentage of UGS. Nevertheless, some limitations need to be considered when interpreting the findings of this study. First, the measurement of UGS was primarily based on the presence of open and accessible green spaces (Lee et al., 2015), which does not account for aspects such as biodiversity and air quality – factors that may have an impact on the health benefits gained from UGS. Given available technology, future studies could consider a broader approach to UGS measurements. The second limitation relates to the small variance of UGS percentage, which as discussed above could explain the lack of association between UGS and stress levels in this study. Follow-up studies in the Philippines could recruit more study sites that would generate a wider range of UGS percentage. Finally, the study utilised a purposive sampling approach which limits the generalisability of the findings. Further work using randomised sampling designs could yield more generalised findings on stress, resilience, and UGS of BPO workers in the Philippines.

## Conclusions

Based on standardised measurements, this study revealed that contact centre agents in the capital region of the Philippines experience high stress levels. The locations of contact centres were shown to have low UGS percentages. After accounting for individual and environment factors, it was found that resilience, household income, and awareness of UGS are significant contributors to the participants’ stress levels. It is recommended that health promotion strategies in contact centres should consider resilience building, enhancing income security, and promoting the awareness of UGS within the workplace vicinity.

## Data Availability

All data may be available upon request from the corresponding author.

## Notes

### Competing Interest Statement

The authors have declared no competing interest.

### Funding Statement

No funding was received for this study

### Author Declarations

University Research Ethics Office, Ateneo de Manila University

